# Design and fabrication of 3D-printed patient-specific soft tissue and bone phantoms for CT imaging

**DOI:** 10.1101/2023.04.17.23288689

**Authors:** Kai Mei, Pouyan Pasyar, Michael Geagan, Leening P. Liu, Nadav Shapira, Grace J. Gang, J. Webster Stayman, Peter B. Noël

## Abstract

The objective of this study is to create patient-specific phantoms for computed tomography (CT) that have realistic image texture and densities, which are critical in evaluating CT performance in clinical settings. The study builds upon a previously presented 3D printing method (PixelPrint) by incorporating soft tissue and bone structures. We converted patient DICOM images directly into 3D printer instructions using PixelPrint and utilized stone-based filament to increase Hounsfield unit (HU) range. Density was modeled by controlling printing speed according to volumetric filament ratio to emulate attenuation profiles. We designed micro-CT phantoms to demonstrate the reproducibility and to determine mapping between filament ratios and HU values on clinical CT systems. Patient phantoms based on clinical cervical spine and knee examinations were manufactured and scanned with a clinical spectral CT scanner. The CT images of the patient-based phantom closely resembled original CT images in texture and contrast. Measured differences between patient and phantom were less than 15 HU for soft tissue and bone marrow. The stone-based filament accurately represented bony tissue structures across different X-ray energies, as measured by spectral CT. In conclusion, this study demonstrated the possibility of extending 3D-printed patient-based phantoms to soft tissue and bone structures while maintaining accurate organ geometry, image texture, and attenuation profiles.

## Introduction

In computed tomography (CT) research and clinical practice, anthropomorphic and geometric phantoms play a crucial role. Highly accurate, customizable, and realistic phantoms are particularly valuable for a variety of purposes, including maintenance, optimization, and development of software and hardware components of scanners. In recent years, there have been significant advancements in three-dimensional (3D) printing technology, resulting in numerous studies on 3D-printed patient-based phantoms for medical imaging [1]–[5]. Compared to conventional phantoms, 3D-printed phantoms are highly accessible, customizable, and cost-effective. For example, inexpensive and widely available fused deposition modeling (FDM) printers can create high-quality anthropomorphic phantoms that accurately depict human anatomy at reasonable costs.

Conventional 3D printing techniques prioritize the replication of object and organ shapes. Typically, these approaches include segmenting organs of interest from CT scans according to their specific densities (HU), converting the results into surface meshes (STL files), 3D-printing each object separately, and then assembling them into a complete phantom [3], [4], [6], [7]. However, each 3D-printed component has a uniform Hounsfield unit (HU), resulting in phantoms with lacking realistic image textures because their HUs cannot be modulated pixel-by-pixel [8]– [11]. Furthermore, the lack of natural transitions between different regions, e.g., organs, leads to loss of detail. A promising alternative is to directly translate digital imaging and communications in medicine (DICOM) image data into G-code. G-code is a Computer Numerical Control (CNC) programming language. G-code instructions tell the printer to move in specific directions and at specific speeds to produce a specific shape or object. One means of controlling the density (as required for CT phantoms) is to vary the filament extrusion rate (per unit time) on a pixel-by-pixel basis while maintaining a constant printing speed. A similar approach was used by Okkalidis et al. [12]–[17]. in conjunction with edge detection and morphological operations to enhance and separate organs. Such processes still yield segmentation errors and loss of small features. Altering the line width by varying the extrusion rate alone does not provide sufficient spatial resolution due to the inherently slow response time of the extrusion process. Our group recently developed PixelPrint [18], a methodology that combines a software tool as well as a standard FDM printer to create phantoms [19]–[23]. In PixelPrint, DICOM images of the original patient are directly converted into G-code on a pixel-by-pixel basis. In order to emulate attenuation at each voxel, density is modeled as a ratio of filament to voxel volume, generating partial volume effects. The filament ratio is continuously modified by varying the printing speed. Polylactic acid (PLA), a common printing filament, allows a print range approximately from -850 to 200 HU at different filament ratios, and has been used to print various patient-based lung phantoms [19].

In parallel, significant progress has been made in developing filament materials suitable for FDM printing in medical applications. Several studies have explored and compared different types of filament materials for printing human soft tissue and bones [24]–[27]. Conventional materials, such as PLA and acrylonitrile butadiene styrene (ABS), are widely available and easy to print with. They have densities ranging from 0.8 to 1.2 g/ml and can represent various human soft tissues for CT or X-ray examinations. Special materials, such as thermoplastic polyurethanes (TPU), can provide distinct physical properties to the print, i.e. durability, strength, and elasticity. Specifically for bone, materials tailored for clinical applications have been introduced for 3D-printed implants. They are biodegradable by the patient’s osteoclasts. As a result, printed objects with such materials can be fused with the patient’s bone, through remodeling during the osteo-cycle [28]– [30]. Additionally, denser PLA filament mixed with gravimetric powdered stone (PLA/Stone) has become commercially available. In previous studies, this type of filament has been utilized for printing phantoms for both diagnostic imaging and radiation therapy [8], [14], [31]–[35]. For printing even higher density objects, commercially available filament materials mixed with micro metal powders, i.e. iron or copper, have also been utilized in phantom studies [6].

This study optimized several aspects of the previously published PixelPrint technique, including filament line spacing and print speed. Furthermore, StoneFil filament, a type of PLA/Stone filament, was utilized to expand the density range of our phantoms in order to print bony structures. Our results illustrate that the PixelPrint technique can create realistic phantoms of the human spine and knee joint with surrounding soft tissue. The resulting phantoms achieved accurate geometry, image texture, and attenuation. Moreover, the presented phantoms exhibited similar spectral attenuation profiles to that of bone structures, which enables their use in various spectral CT applications.

## Materials and Methods

### 2.1 PixelPrint and 3D printing

The previously published PixelPrint algorithm was used to create G-code from CT image data to produce 3D-printed phantoms [18]. Briefly, density information was extracted from the clinical patient images to generate filament lines that varied in width according to the HU of individual pixels. These lines were uniformly spaced within each layer and perpendicular on adjacent layers. By adjusting the filament line widths pixel-by-pixel, volumetric filament per unit space, or infill ratio, was varied despite only using one type of filament. These different infill ratios then produced different attenuation in CT images due to the partial volume effect.

In this study, the filament lines were equally spaced at 0.5 mm. The width of the filament line changed at resolution of 0.167 mm. The minimum and maximum line widths were 0.2 and 0.5 mm, corresponding to the infill ratio ranging between 40% and 100%, respectively. Keeping a constant extrusion rate, the print head traveled at varying speeds based on the width of the extruded filament line. The slowest speed was 180 mm/min for the widest width of 0.5 mm, while the fastest was 450 mm/min for the smallest width of 0.2 mm. Each layer had a uniform height of 0.2 mm. The resulting volumetric rate of filament extrusion during the whole print remained constant at 18 mm^3^/min. To prevent overlapping of lines in consecutive layers with the same filament line direction, an offset of 0.167 mm (1/3 of the 0.5 mm line spacing) was introduced.

All phantoms were printed with Lulzbot TAZ 6 or Sidekick 747 (Fargo Additive Manufacturing Equipment 3D, LLC Fargo, ND, USA), paired with M175 v2 tool heads and 0.40 mm steel nozzles. StoneFil filament (FormFutura, AM Nijmegen, the Netherlands) with a diameter 1.75 mm was utilized. The temperature of the nozzle was set at 200 °C and the bed was warmed to 50 °C to enhance adherence. Acceleration of the print head was to 500 mm/s^2^ and the threshold (jerk setting) was 8 mm/s.

### 2.2 Phantom design

#### Micro-CT phantom

Three cylindrical phantoms were designed and produced using PixelPrint filament lines to examine their stability and reproducibility. These filament lines constructed a matrix smaller than the typical resolution limit of clinical CT scanners. Three phantoms were printed with identical G-code instructions. These phantoms are 60 mm in length and 20 mm in diameter. Each of them consists of four sections with different but homogeneous infill ratios (100%, 70%, 50% and 30%). StoneFil filament lines were printed at a spacing of 1 mm in all four sections but with corresponding line widths of 1.0, 0.7, 0.5, and 0.3 mm, respectively. A thin outer layer was added to the phantom for support, particularly for low infill ratio sections.

#### Calibration phantom

To compute the conversion between StoneFil filament infill ratios and HUs, a calibration phantom was designed. The phantom is a cylinder with a diameter of 10 cm and height of 1 cm. It consists of seven equally divided pie slice-shaped sections. Each section was printed at a fixed line spacing of 0.5 mm but with different filament line widths (0.2 - 0.5 mm), corresponding to seven infill ratios (40 -100%, with 10% intervals).

#### Cervical vertebrae phantom

Institutional Review Board (IRB) approved this retrospective study. A cervical vertebrae phantom was created based on a patient image volume (10 × 10 × 10 cm^3^) that was acquired on a clinical CT scanner (Siemens SOMATOM Definition Edge, Siemens Healthcare GmbH, Erlangen, Germany) at a tube voltage of 120 kVp with a standard diagnostic protocol. Table 1 lists detailed acquisition and reconstruction parameters for the patient scan. The patient data consist of four cervical vertebrae (C4 to C7), including the trachea and esophagus. A circular region of interest with a diameter of 10 cm was cropped in axial slices to form the phantom. HUs were converted to infill ratios based on the calibration phantom.

**Table 1.** Acquisition parameters of CT image for phantom generation.

|  | *Cervical vertebrae* | *Knee* |
| --- | --- | --- |
| Scanner model | Siemens SOMATOM Definition Edge | Philips iQon Spectral CT |
| Tube voltage | 120 kVp | 120 kVp |
| Tube current | 105 mA | 196 mA |
| Rotation time | 1000 ms | 1026 ms |
| Spiral pitch factor | 0.8 | Axial |
| Exposure | 131 mAs | 201 mAs |
| CTDI<sub>vol</sub> | 8.85 mGy | 17.1 mGy |
| Collimation width | 0.6 / 38.4 mm | 0.625 / 40.0 mm |
| Slice thickness | 0.6 mm | 0.67 mm |
| Reconstruction filter | I26s\3 | B |
| Field of view | 99.75 x 99.75 mm<sup>2</sup> | 304 x 304 mm<sup>2</sup> |
| Matrix size | 228 x 228 pixel<sup>2</sup> | 512 x 512 pixel<sup>2</sup> |
| Pixel spacing | 0.4375 mm | 0.5938 mm |

Collimation width values are noted as single / total collimation width.

#### Knee phantom

A knee phantom was similarly generated using a patient scan on a clinical dual-layer CT scanner (IQon spectral CT, Philips Healthcare, the Netherlands) at a tube voltage of 120 kVp, as detailed in Table 1. A circular region of interest with a diameter of 10 cm was cropped from the axial slices of the patient’s left knee. HUs were then converted to infill ratios.

### 2.3 Data acquisition

Three micro-CT phantoms were separately scanned on a commercial micro-CT (U-CT system, MILabs, CD Houten, the Netherlands) with a tube voltage of 50 kVp. In addition, these phantoms were also scanned on a clinical dual-layer CT system (IQon spectral CT, Philips Healthcare, the Netherlands) at a tube voltage of 120 kVp with a high-resolution protocol and a small field-of-view of 100 mm. Additional acquisition and reconstruction parameters of the two scans are listed in Table 2. Micro-CT images were exported from the scanner and reprocessed with a multi-planar reconstruction algorithm (MPR) in Horos (Horos Project, Annapolis, MD, USA) to ensure filament lines were parallel to the axial plane.

**Table 2.** Scan protocols for the micro-CT phantom.

|  | Micro-CT | Clinical CT |
| --- | --- | --- |
| Scanner model | MILabs U-CT | Philips IQon Spectral CT |
| Tube voltage | 50 kVp | 120 kVp |
| Tube current | 0.21 mA | 130 mA |
| Rotation time | 54 s | 1.923 s |
| Spiral pitch factor | Axial scan | 0.39 |
| Exposure | 11.3 mAs | 250 mAs |
| CTDI<sub>vol</sub> | 69 mGy | 16.4 mGy |
| Collimation width | - | 0.625 / 40.0 mm |
| Slice thickness | 0.08 mm | 0.67 mm |
| Reconstruction filter | - | YC |
| Field of view | 22.16 x 22.16 mm<sup>2</sup> | 100 x 100 mm<sup>2</sup> |
| Matrix size | 277 x 277 pixel<sup>2</sup> | 512 x 512 pixel<sup>2</sup> |
| Pixel spacing | 0.080 mm | 0.195 mm |

Collimation width values are noted as single / total collimation width.

The calibration, cervical vertebrae, and the knee phantom were scanned inside the QRM chest phantom (Quality Assurance in Radiology and Medicine GmbH, Möhrendorf, Germany) with the clinical dual-layer CT system. Protocol parameters matched those of the original clinical examination of the patient, with the same pixel spacing and slice thickness in Table 1. For the cervical phantom, a 400 mg/ml QRM hydroxyapatite (HA) insert was additionally scanned with the phantom as a reference for bone mineral density. For both patient-based phantoms, additional high dose scans were performed at 1000 mAs while keeping the other scanning parameters the same. This high exposure scan was included to reduce noise for image quality comparisons.

### 2.4 Calibration and data analysis

For computing the conversion between HUs and infill ratios, mean and standard deviation HU values of seven areas were measured in the calibration phantom. Square regions of interest (ROI) of 19 × 19 pixel^2^ (13 × 13 mm^2^) were manually placed in each of the seven density regions within 10-mm-thick center of the phantom. A linear regression was computed, and the resulting Pearson’s correlation coefficient (r) was reported. All measurements were performed on a workstation with ImageJ (U. S. National Institutes of Health, https://imagej.nih.gov), and all analyses were computed with Python (Python Software Foundation, https://www.python.org/).

For the cervical vertebrae phantom and the knee phantom, CT images were exported from the scanner and registered to the original patient data (2D-wise) using the OpenCV Library (Open Source Computer Vision Library [36], https://opencv.org). Mean and standard deviation in regions of interest for different tissue types were measured. Line profiles of the phantom scan were also compared with the original patient scan. Additionally, virtual monoenergetic images from 40 to 200 keV were extracted to quantify the spectral response of the bone regions within the patient-based phantoms.

## Results

The high reproducibility of PixelPrint was demonstrated by comparing three identically manufactured phantoms (Figure 1). In micro-CT scans of the phantoms, the grid-like structures generated by PixelPrint were clearly visible. Filament lines printed within each region had equal spacings of 1 mm and a constant width in all three phantoms in the micro-CT scans. The layered structure with introduced offsets (1/3 of 1 mm line spacing) was distinctly visible in orthogonal views (Figure 1f, 1g, 1h). However, in clinical CT scans with high resolution protocols, these structures were imperceptible because their size was smaller than the detector resolution. Instead, they appeared as constant regions due to partial volume effect (Figure 1e). Furthermore, both the micro-CT and clinical CT scans showed a high linear relationship between infill ratios and mean HUs in four regions (Pearson’s correlation coefficient *r* = 0.984 and 0.982, respectively).

**Figure 1.**
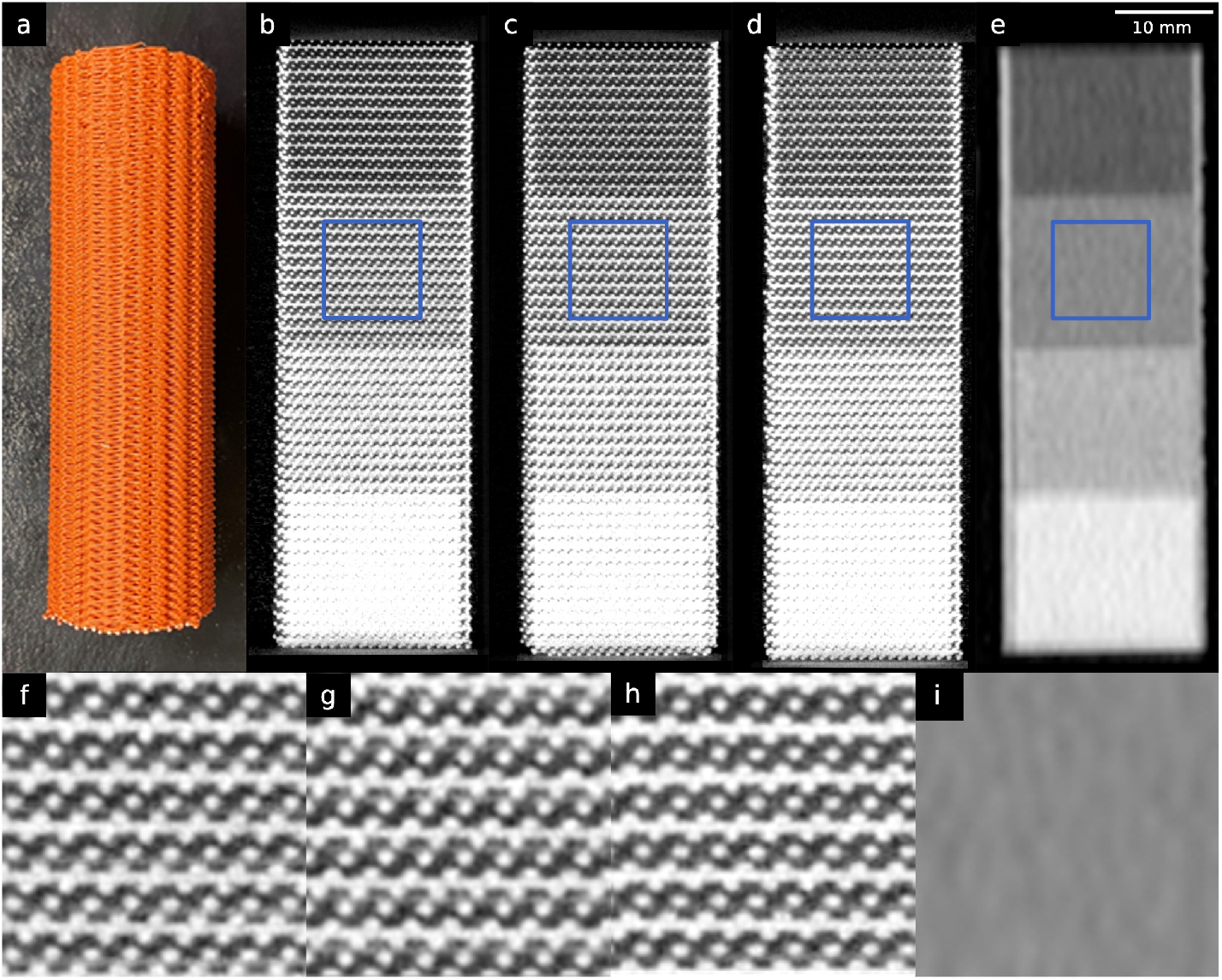
Micro-CT phantoms. (a) A photo of one of the three printed micro-CT phantoms. (b)-(d) Orthogonal views of the three different micro-CT phantoms scanned on a micro-CT. (e) Clinical CT image of one of the micro-CT phantoms. (f) – (i) Zoomed views of the regions enclosed by blue squares in (b) - (e). Window level/width are - 750/3500 HU for micro-CT images and 0/2000 HU for clinical CT images.

In the calibration phantom, the infill ratio and HU also demonstrated excellent linearity across the seven regions (Figure 2). The highest infill ratio (100%) region measured 851 ± 24.7 HU, while the lowest infill ratio (40%) measured -227 ± 25.4 HU. Pearson’s correlation coefficient of greater than 0.99 indicated a very high positive linear correlation between infill ratios and HUs. A conversion equation was computed for converting HU to infill ratio:

**Figure 2.**
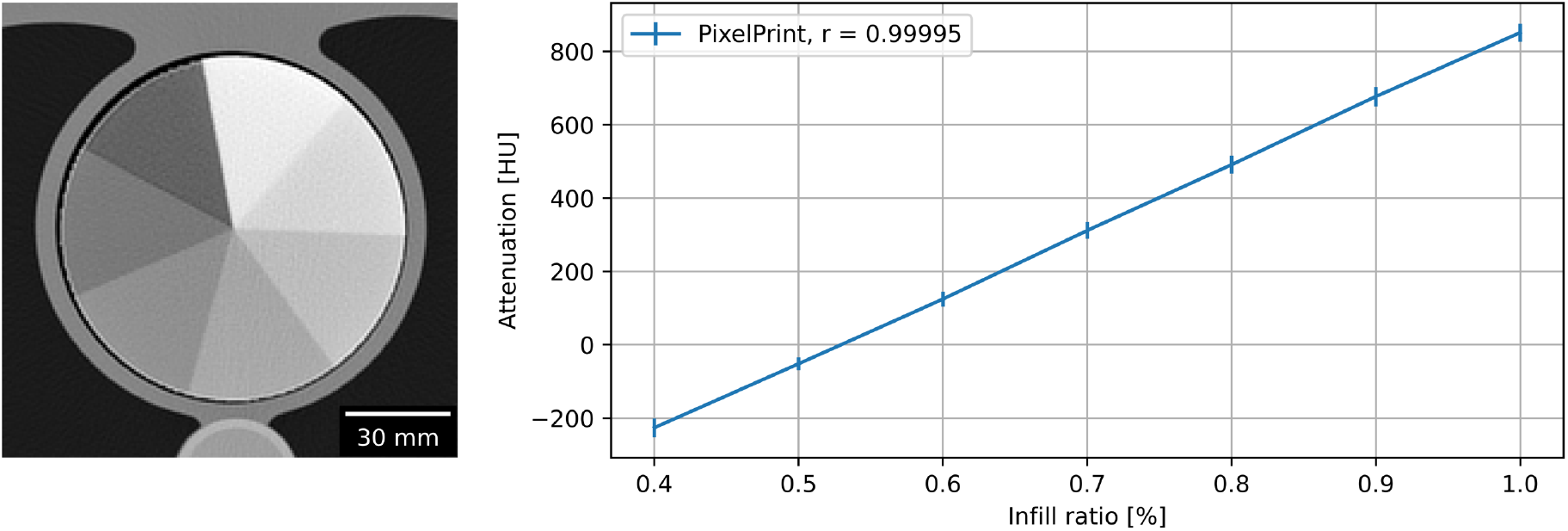
Linear correlation of filament infill ratio and HUs. (a) CT image of calibration phantom. Window level and width are 0 HU and 2000 HU. (b) Linear relationship between attenuation and infill ratio. Mean and standard deviation were measured in regions of interest in each area with a distinct infill ratio. Standard deviations are indicated with error bars.

Figure 3 shows the CT images of the cervical vertebrae phantom, while Figure 4 illustrates the images of the knee phantom. The PixelPrint phantoms closely resembled the original CT images in terms of contrast and features, with both the shape and the details inside the bones very well reproduced. The image features in the high dose knee phantom scan appeared as sharp as the original patient image.

**Figure 3.**
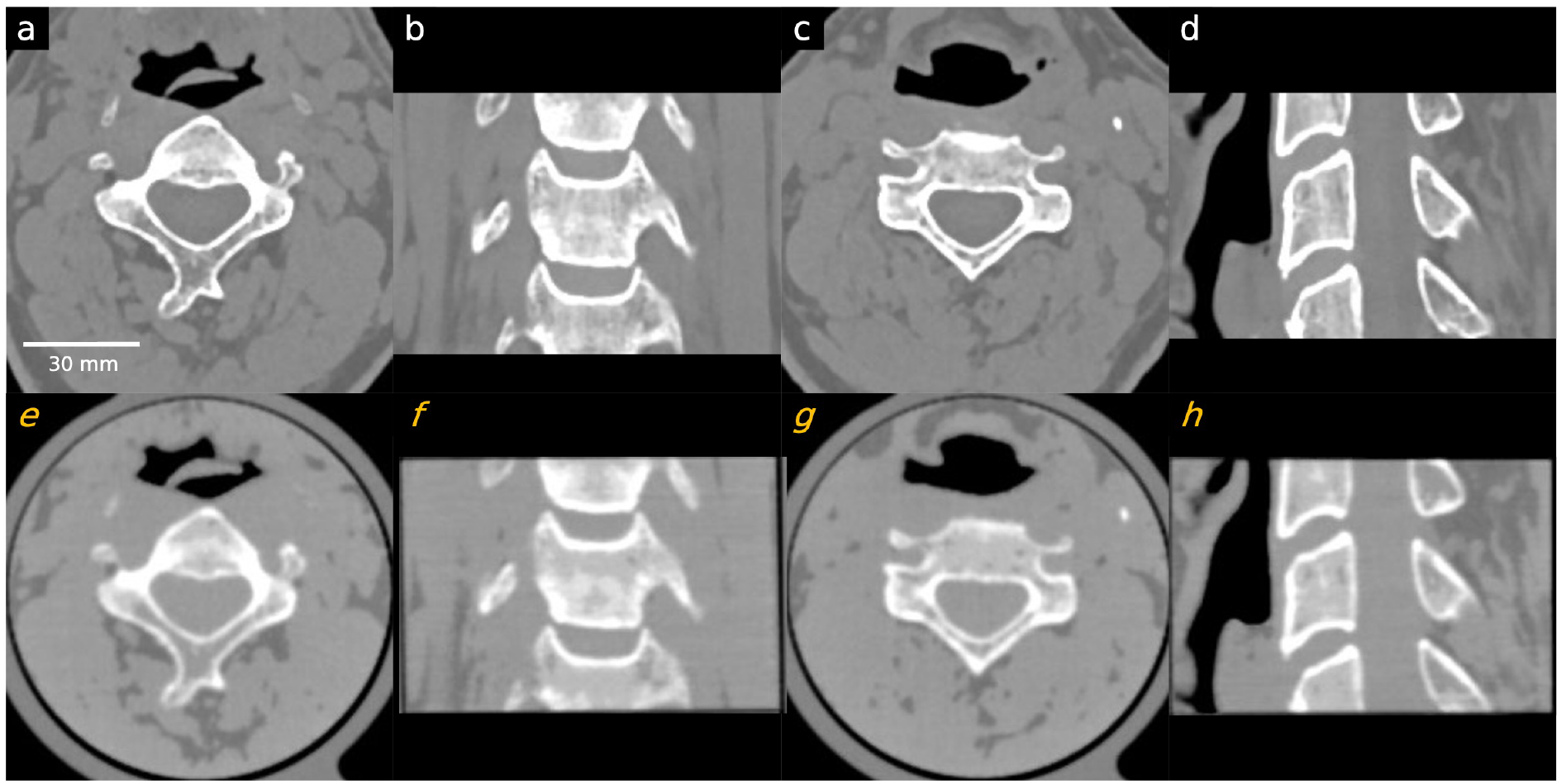
Comparison between patient CT images and the PixelPrint cervical phantom images. Images in the first row (a-d) are original DICOM images used to create the PixelPrint cervical phantom. Images on the second row (e-h) are the CT images of the phantom. All images have window level of 0 HU and width of 1200 HU. Sagittal and coronal images are not registered but are approximately at the same location.

**Figure 4.**
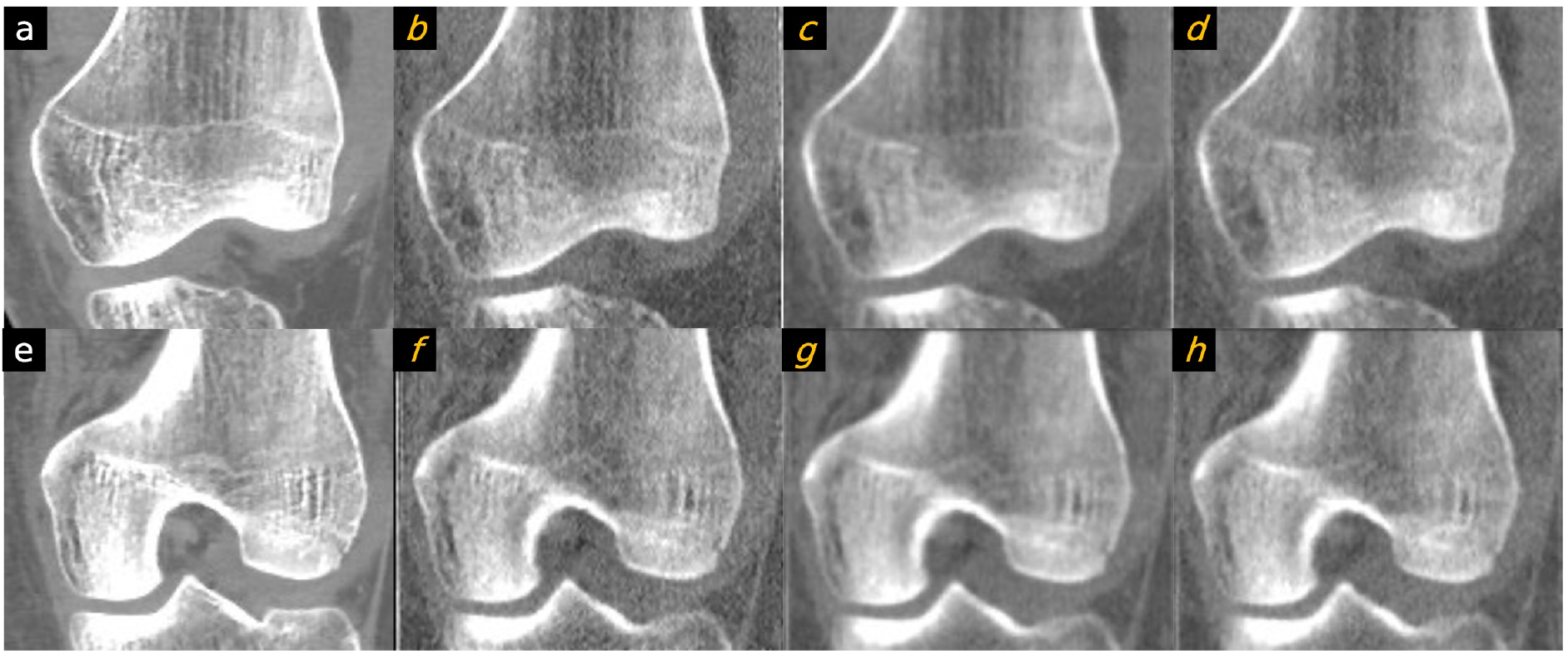
Comparison between patient image and the PixelPrint knee phantom. Images in the first column are original DICOM images used to create the PixelPrint knee phantom. Images on the second to fourth column are the CT images of the phantom: (b/f) high dose sharp kernel. (c/f) high dose standard kernel. (d/h) standard dose sharp kernel. All images have window level of 0 HU and width of 1200 HU. Images are not registered but are approximately at the same location.

Patient phantoms showed high accuracy. Line profiles indicated a match in HUs between the CT image of the cervical vertebrae phantom and the patient data (Figure 5). Quantitative measurements in selected regions of trabecular and cortical bones, as well as adipose- and muscle-like soft tissues, are provided in Table 3. Measurements indicated that, except for the cortical bone, all other regions had differences of less than 15 HU compared to the patient image. Due to the density limitations of the utilized filament, HUs for the cortical bone (region 3 in Figure 6) were lower than expected.

**Figure 5.**
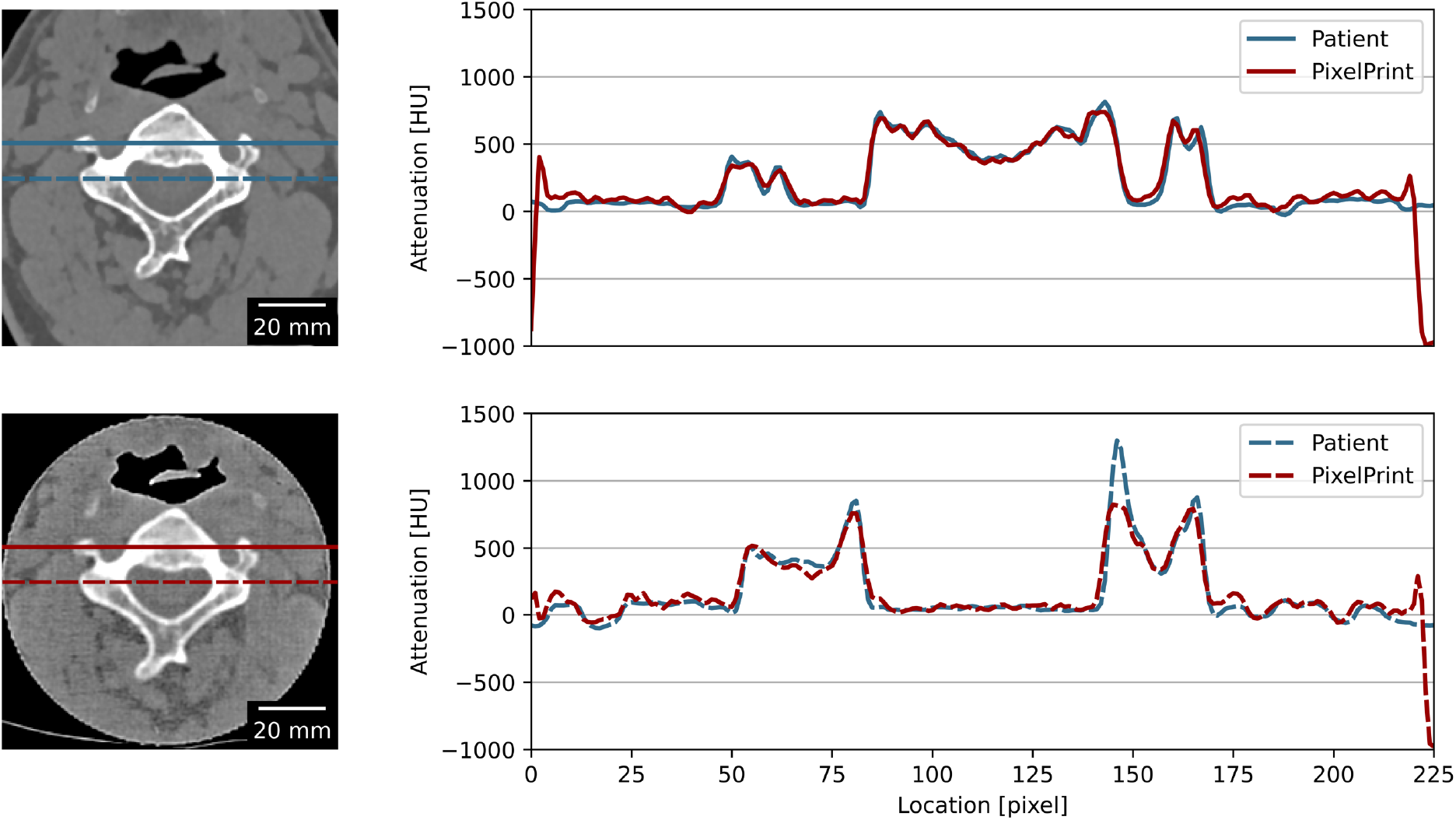
Line profiles of the PixelPrint phantom and the patient CT images. Images on the left show the CT images of the phantom (upper) and the patient images (lower). Red and blue lines indicate the location used to measure the line profile plot on the right. Window level and width are 0 HU and 2000 HU. Images were assumed to be at the same location and registered 2D-wise.

**Table 3.** Measured Hounsfield units for different tissue types in patient and phantom.

| Area |  | Patient |  |  | PixelPrint Phantom |  |  | difference |
| --- | --- | --- | --- | --- | --- | --- | --- | --- |
| | | mean ± stdev | min | max | mean ± stdev | min | max | |
| 1 | Bone I | 49.3 ± 31.2 | 3.4 | 175.8 | 57.1 ± 45.5 | -45.0 | 185.3 | +7.8 |
| 2 | Bone II | 363.9 ± 50.9 | 242.2 | 452.1 | 349.2 ± 47.0 | 232.8 | 504.6 | -14.7 |
| 3 | Cortical Bone | 1319.7 ± 87.4 | 1008.9 | 1406.8 | 800.6 ± 14.5 | 760.6 | 837.9 | -519.0 |
| 4 | Soft tissue I | 55.6 ± 8.5 | 36.5 | 76.3 | 53.1 ± 24.1 | 5.4 | 107.3 | -2.5 |
| 5 | Soft tissue II | -78.1 ± 13.5 | -105.9 | -55.8 | -66.2 ± 44.7 | -174.9 | 9.6 | +11.9 |

All measurements are in HUs. Stdev stands for standard deviation. Patient and phantom images were assumed to be the same z location and registered 2D-wise.

**Figure 6.**
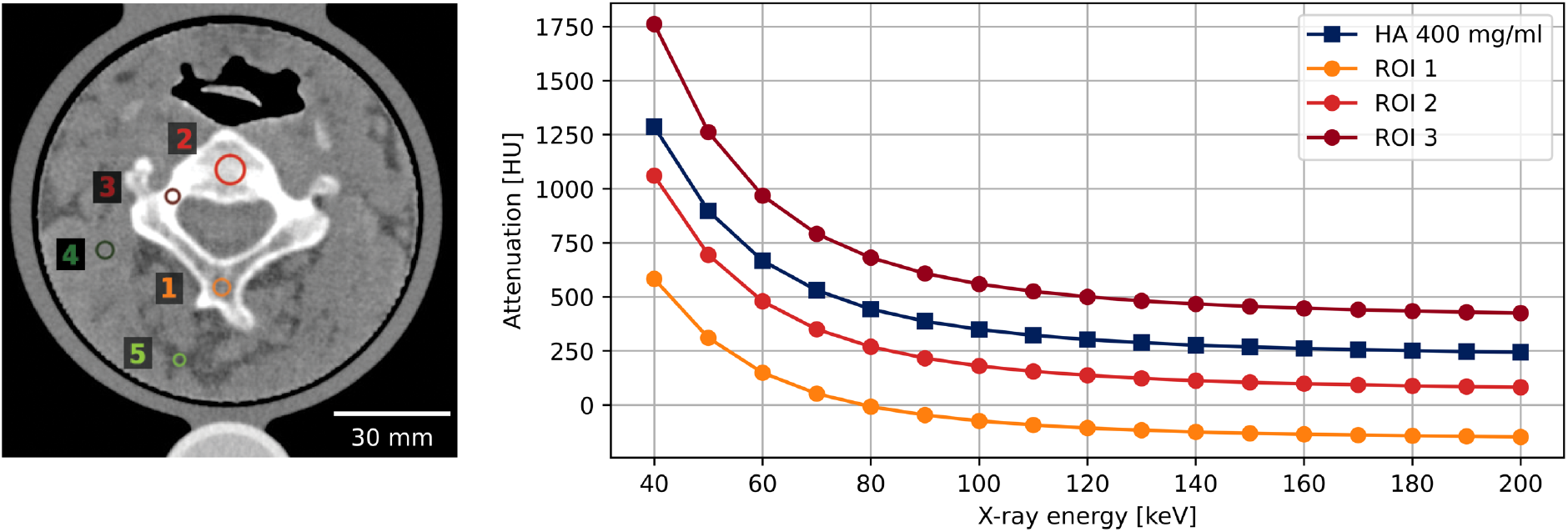
Virtual monoenergetic HU measured with spectral CT. Regions of interest (ROI) are marked in left. Window level and width are 100 and 800 HU. Reference values from a 400 mg/ml hydroxyapatite (HA) insert are marked by dark blue squares.

Comparable spectral characteristics of the phantom to those of human bone were observed. Figure 6 depicts the spectral attenuation profile of various regions of interest (marked in the left panel) and a 400 mg/ml hydroxyapatite insert (displayed in dark blue in right panel). It is noteworthy that the phantom was fabricated using only one type of filament, and thus, the background, which represents soft tissue, has artificial amounts of calcium.

## Discussion

This paper demonstrated how PixelPrint can be utilized to create patient-specific 3D printed bone and soft tissue CT phantoms using one filament. Our approach provides economical and efficient means of producing high resolution CT phantoms, exhibiting excellent accuracy in HU and image texture characteristics in CT scans. These phantoms are useful for a wide range of academic research and clinical evaluation of CT performance.

In contrast to prior studies of image-based 3D printed bone phantoms using slices of the human head/skull [13], chest/thoracic cage [15], pelvis [14] and femoral shaft [6], this study printed the human cervical vertebrae with surrounding soft tissue. Human vertebrae particularly present a challenging task for 3D printing, as they contain intricate details and are comparatively smaller in size. Nevertheless, these areas, especially in combination with the adjacent tissues, are not only fundamental in clinical diagnostic applications, such as the assessment of severe fractures or degenerative diseases, but also crucial in surgical interventional planning. Our phantoms possess the potential to be utilized for those applications, such as optimizing CT protocols for the assessment of bone mineral density [37] among others. Here, only human cervical vertebrae and knee joint phantoms were printed, but the approach can be extended to any bone structure. With StoneFil filament, a range of approximately -227 HU to 851 HU for a CT scans with a tube voltage of 120 kVp can be reliably printed using PixelPrint, with a deviation of less than 15 HU compared to patient data. This range covers most tissue types in the human body and is applicable to various research applications.

Continuing our previously published research on the PixelPrint lung phantom [18], [19], this study not only extended the types of human tissue printed, but also enhanced the resolution and stability of PixelPrint. Filament line spacing was reduced from 1.0 to 0.5 mm, potentially doubling the resolution capabilities of the printed phantoms. Phantoms produced using this approach can have greater filament coverage and finer details in a given area, serving as valuable tools to evaluate the efficacy of novel higher resolution CT systems such as photon-counting CT [38]–[40]. Printing finer lines with PLA/Stone filament poses more challenges to printer stability control and requires finer system tuning. By optimizing extrusion rate, printing speed, nozzle temperature, and acceleration speed, PixelPrint can still produce highly accurate patient phantoms in reliable stability as demonstrated by qualitative and quantitative evaluation. Additionally, micro-CT acquisitions revealed that filament lines and underlying structure can be generated with high degree of consistency.

With the growing popularity and accessibility of 3D printing technology, a variety of printing filaments are now available for printing human bone and soft tissue. Several studies have discussed materials for 3D-printed phantoms in CT [24]–[26]. Novel filament materials composed of hydroxyapatite and biocompatible, biodegradable polymers, such as CT-Bone (Xilloc Medical Int., Sittard-Geleen, the Netherlands), can be utilized for printing synthetic bone implants that rapidly induce bone regeneration and growth [41], [42]. Filaments made from composites of fatty acids and ceramic powders have also been explored [28]. However, bone-like filaments available in the general market (FibreTuff, Toledo, OH, USA), suitable for medical surgery purposes [29], [30], do not necessarily have high radiometric densities and are not capable to reach much higher than 400 HU in CT scans. While cancellous bone is only about 300 to 400 HU in CT images, cortical bone can range from 500 HU and up to over 1900 HU [43]. By contrast, materials such as vinyl and PLA with stone (PLA/stone) can offer up to nearly 1000 HU at 96.9% infill ratio at tube voltage of 120 kVp, as they exhibit relatively higher X-ray absorption. Additionally, considering materials for spectral CT phantoms, high impact polystyrene (HIPS) based filaments may be suitable for mimicking CT numbers in applications where energy dependence is important [26], because they show similar spectral profiles as the human body. In this study, we employed StoneFil filament, one type of PLA/stone filament. Unlike normal PLA, StoneFil filament is gravimetrically filled with 50% powdered stones, resulting in significantly higher material density and enabling denser printed objects. Carbonate calcium-containing limestones exhibit a similar X-ray response in CT to that of human bone, whose density can be attributed to hydroxyapatite. This property was reflected in the the spectral response of the printed vertebrae with its similarity to that of hydroxyapatite.

This study has a few limitations: (i) The filament used in our study did not encompass the entire range of Hounsfield Units (HU) required for bone structures. Future research should focus on the development of next-generation filaments that cover the full HU range while preserving spectral capabilities. (ii) The calcium-based material used in the printing process was applied to the entire print, including soft tissue regions. While this approach does not severely impact performance in conventional CT applications, it may have an influence on the evaluation with spectral CT. To achieve the full dynamic range with spectral characterization for both soft tissue and bone, further development of multiple print head systems will be required. (iii) The printed phantoms were limited to a specific field of view. Future studies should explore the potential to print larger anatomical regions, such as the entire chest or abdomen.

## Conclusion

Our study successfully showed the feasibility of using PixelPrint and stone-based filament to 3D-print patient-based bone phantoms with surrounding soft tissue for use in clinical CT applications. The resulting phantoms accurately replicated patient’s CT imaging, including precise organ geometry, image texture, and attenuation profiles for spectral CT, which can greatly benefit both academic research and clinical applications.

## Data Availability

All data produced in the present study are available upon reasonable request to the authors.

## Acknowledgements

We acknowledge support through the National Institutes of Health (R01CA249538, R01EB030494, and R01EB031592).

## Author Contribution

K.M., M.G. and P.N. devised the project, the main conceptual ideas and proof outline. N.S., G.G and J.W. contributed to the design and the research.

K.M. implemented the idea. K.M. P.P. and L.L. performed the experiments and measurements.

K.M. analysed the data and wrote the manuscript with support from L.L. and P.N. N.S., G.G and J.W. helped the revision of the manuscript.

## Additional Information

The authors have no relevant conflicts of interest to disclose.

## Data Availability

Datasets generated during this study are available from the corresponding author upon reasonable request.

